# Improved Data Alignment Between National HIV Reporting Systems, Mozambique, 2017–2023

**DOI:** 10.1101/2024.11.19.24317535

**Authors:** Orrin Tiberi, Lindsay Templin, Ferreira Ferreira, Herminio Nhaguiombe, Aleny Couto, Joe Lara, Ryan Keating, Jose Mizela, Lucio Matsimbe, Helio Magaia, Morais da Cunha

**Affiliations:** National STI and HIV/AIDS Control Program, Ministry of Health Mozambique; Division of Global HIV & TB (DGHT), U.S. Centers for Disease Control and Prevention (CDC), Mozambique, Maputo; U.S. Agency for International Development (USAID), Maputo, Mozambique

**Keywords:** HIV, Data Quality, Data Alignment, PEPFAR, ART

## Abstract

**Introduction:** Data quality improvements have aimed to align national reporting systems for Ministry of Health (MISAU) and PEPFAR. For example, in 2019 the patient Master Card improved systematic data collection process and sites were selected for intensified support including data quality activities (AJUDA). This study aims to compare reported data between MISAU and PEPAFR to understand data alignment trends.

**Methods:** The annual number of PLHIV on ART, newly initiating ART and HIV status in first ANC visit as reported by MISAU and PEPFAR were compared for 2017-2023. The absolute difference was calculated as MISAU-reported number minus PEPFAR-reported number; proportional absolute difference as the absolute difference over the MISAU-reported number and assessed the slope of the difference over time.

**Results:** During 2017–2023, median absolute difference for PLHIV on ART was 104,940 (range = 154,901 [2018] to 1,598 [2020]), and median proportional difference was 5.0% (range = 0.1% [2020] to 12.8% [2018]), with a trend towards improved concordance. A similar trend was found in positive HIV status at first ANC (10.4% [2017] to 0.0% [2023]), as well as with newly initiated on ART (2.7% [2017] to 0.3% [2023]).

**Conclusion:** In the three indicators analyzed, there were improvements in data alignment between the years of 2017 and 2023, with increased alignment in different years and for different reasons. Continued improvements will support programming and increase certainty for tracking progress toward the UNAIDS 95-95-95 goals to end HIV.

## Introduction

Among implementing agencies aligned in a shared vision, a common understanding of the HIV response based on high quality data allows for a unified approach to programming and goal setting. The Joint United Nations Programme on HIV/AIDS (UNAIDS) Global AIDS Strategy 2021–2026 includes data as one of three critical cross-cutting areas that are essential to inform and guide the HIV response.^1^

The Mozambique Ministry of Health (MISAU) relies on manual aggregation of patient records from registers for facility level program monitoring and official reporting in a national reporting platform. The U.S. President’s Emergency Plan for AIDS Relief (PEPFAR) has implemented an electronic medical record (EMR) system based in OpenMRS to capture patient-level data and electronically generate facility-level indicators, which are submitted quarterly to PEPFAR. Electronic systems, such as OpenMRS, are used to improve the management of services, and have been presented as a useful tool to improve data quality and clinical follow-up of users.^2,3^ However, there is no perfect system, and some challenges identified for electronic systems include data completeness, interoperability, inconsistencies, over-reporting of users, changes in the definition of indicators, and differences in collection systems, among others.^4-6^

Since 2017, various activities focused on data quality improvements aimed to align MISAU- and PEPFAR-reported data (Figure 1). In 2017 a national data alignment activity was initiated, which focused on the annual reconciliation of site-level results between MISAU and PEPFAR.^7^ In 2019, a new patient chart was launched, which required a recount of the antiretroviral therapy (ART) archive and allowed for a common base between the EMR and the MISAU reporting.^8^ In 2020, PEPFAR finalized their pivot to higher burden health facilities, including enhanced data quality improvement activities, impacting data quality for both PEPFAR and MISAU.^9^

**Figure 1:**
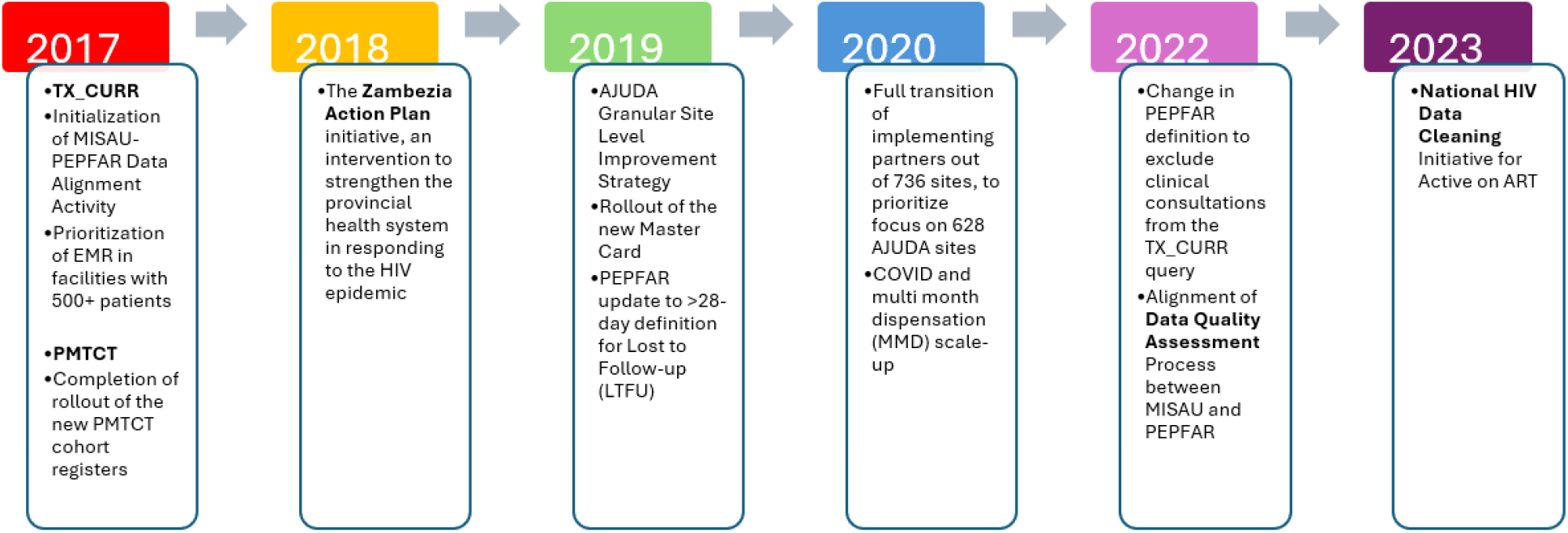
Activities That Contributed to Increased Alignment Between MISAU and PEPFAR from 2017 to 2023

Changes to reporting and indicator definitions have also been introduced to increase data alignment. In 2023, PEPFAR further updated its definition of people living with HIV (PLHIV) active on ART to be exclusively based on the ARV drug pickups, in alignment with the MISAU definition. In addition, the continued improvements in interoperability and standardization of the EMR system, covering 83% of PLHIV on ART in 2017 and 85% in 2023, has contributed to improved data quality.^10^

To understand the impact of these data improvement and alignment activities, we compared data reported for key indicators through the MISAU with PEPFAR national HIV data reporting systems.

## Methods

We compared the calendar year totals for results reported by MISAU in the yearly reports and those reported by PEPFAR for three key program indicators: PLHIV active on ART, newly initiated on ART, and HIV-positive status at first antenatal care (ANC) visit.^11^

For PLHIV active on ART, the MISAU definition counts clients with ≤59 days since last missed antiretroviral (ARV) drug pick-up; the PEPFAR definition counts clients with ≤28 days. Prior to October 2019, the PEPFAR definition was ≤90 days since last ARV drug pick-up or of clinical consultations. Comparisons of PEPFAR and MISAU report based on the EMR system suggest an expected difference of 0-2% depending on patient fluctuations. During 2017–2023, MISAU and PEPFAR defined newly-initiated on ART as a PLHIV with their first ARV drug pick-up during the data reporting period. Finally, HIV-positive status at first ANC visit measures the number of pregnant women who are living with HIV with known status as well as those newly diagnosed with HIV infection at first ANC. MISAU reports these data at the end of a 6-month cohort, while PEPFAR reports on women in their first ANC within the 3-month reporting period. For this analysis, cohorts were aligned for comparability.

For each indicator, we calculated the absolute and percent difference between MISAU and PEPFAR reported results per calendar year with MISAU results as the baseline. We used publicly available aggregate monthly national-level MISAU results, and quarterly results reported to PEPFAR.^a^ Data analysis was carried out in Excel v2409, where differences between the data were calculated in absolute numbers and percentages.

## Results

For PLHIV active on ART, the median difference was 104,940 (range = 1,598 [2020] to 154,901 [2018]), and median proportional difference was 5.0% (range = −0.1% [2020] to 12.8% [2018]). The proportional difference decreased from 2017 (11.5%) to 2023 (5.0%).

For PLHIV newly initiated on ART, median difference was 2,297, ranging from 9,078 in 2017 to 48 in 2022. The percent difference was <2.0% during 2017–2023, except 2017 (2.7%), with the lowest percent difference in 2022 (<0.01%) as seen in Figure 2.

**Figure 2.**
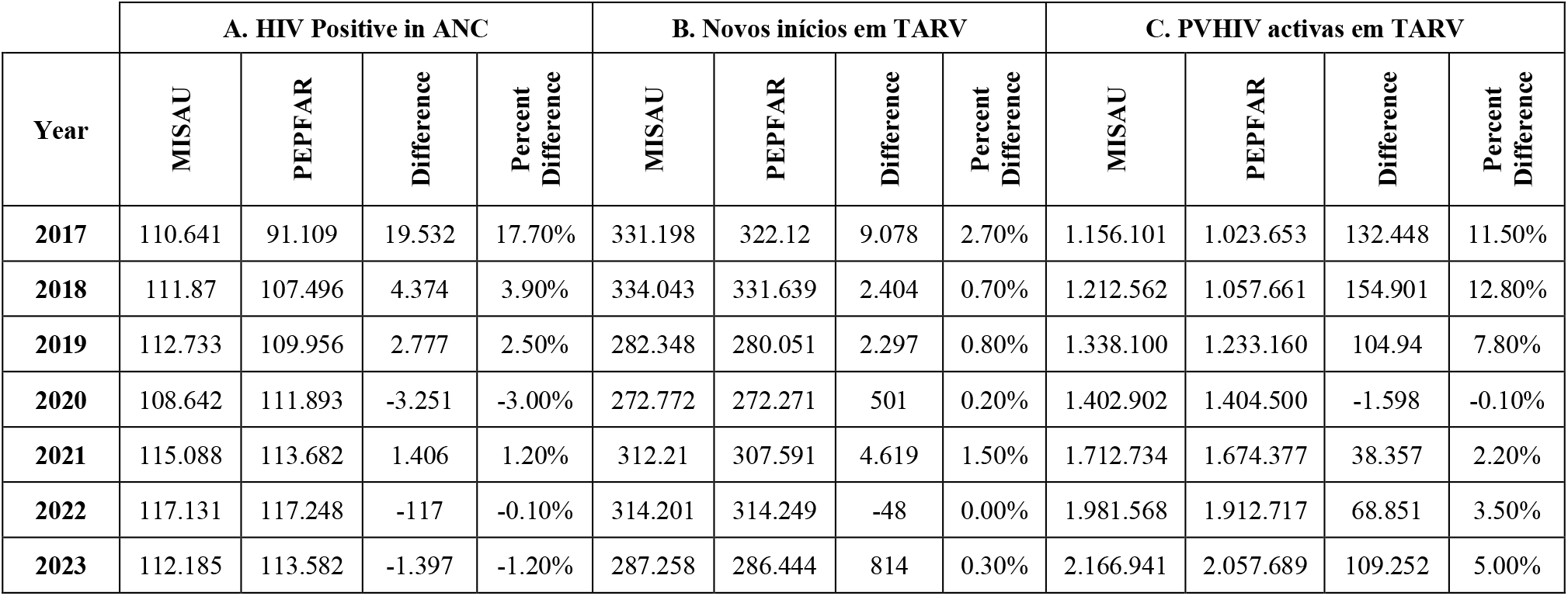
Trends of Alignment for the Three Key Indicators between MISAU and PEPFAR

For HIV-positive status at first ANC, the absolute and proportional difference decreased from 2017 (absolute difference = 19,532; proportional difference = 17.7%) to 2023 (absolute difference = 36; proportional difference = <0.01%), moving towards almost complete alignment as seen in Figure 2 graphic C. The median absolute difference during the analysis period was 1,842, and 3 years had a proportional difference >2.0% (2017, 2018, 2020).

## Discussion

There are differences in the impact of the interventions carried out between 2017 and 2023 to improve the quality and alignment of data between MISAU and PEPFAR. For the indicator active on ART, the large impact of an intervention for data improvement was the change to the new care and treatment instruments in 2020, where the absolute difference dropped from 7.8% to 0.1%. The change can be attributed to the fact that all the US recounted the ART file. For newly initiating on ART, it was between 2017 and 2018, when the country introduced the quarterly data alignment activity between the two systems. In addition, this indicator was key to the test-and-start approach in 2018, which could have been another reason for this alignment. Finally, for the HIV-positive pregnant woman indicator from the ANC, the package of new instruments that was introduced in mid-2017 included reinforcement for data completion and reporting flow and resulted in improved alignment.

Without aligning data definitions, reporting periods, and reporting structures, complete data alignment will remain a challenge. The impact can be seen in the discrepancy for PLHIV on ART between 2020 and 2023, where a combination of differences in reporting methods, periods, and data quality issues led to an increase in proportional differences between the two systems. Despite these challenges, there are new initiatives to improve data alignment, such as the unified approach to the annual national data quality assessments and a national ART chart cleaning. Initial results from the first quarter of 2024 reflected improved data quality, decreasing the absolute difference of PLHIV active on ART substantially by March 2024.

## Conclusions

Since 2017, the MoH and PEPFAR have improved data alignment for three key HIV indicators, despite differences in HIV reporting systems, which allows for greater collaboration in data analysis and quality reviews. These ongoing improvements will support programming and increase security to track progress towards the UNAIDS 95-95-95 targets to end HIV in Mozambique. Finally, with the upcoming revision of the ART patient file and reporting flows there is an opportunity to build on the success of previous improvements in data quality that would allow for sustained improvements in data.

## Data Availability

All data produced are available online from the PEPFAR Monitoring, Evaluation, and Reporting (MER) Database or from the publicly available Ministry of Health HIV Annual Reports

https://mer.amfar.org/

https://old.misau.gov.mz/index.php/its-hiv-sida

## Acknowledgements

We would like to acknowledge the support of PEPFAR Mozambique and the Mozambican Ministry of Health – National Public Health Directorate in the conception, elaboration, and approval of this short report.

## Funding Acknowledgement

This manuscript has been supported by the President’s Emergency Plan for AIDS Relief (PEPFAR), through the Centers for Disease Control and Prevention (CDC), The findings and conclusions in this manuscript are those of the author(s) and do not necessarily represent the official position of the funding agencies.

## Footnotes

a This activity was reviewed by CDC, deemed not research, and was conducted consistent with applicable federal law and CDC policy. See e.g., 45 C.F.R. part 46.102(l)(2), 21 C.F.R. part 56; 42 U.S.C. §241(d); 5 U.S.C. §552a; 44 U.S.C. §3501 et seq.

